# The Age-Varying Association Between BMI and Height-for-Age in Vietnamese Schoolchildren: A Sex-Stratified Quantile Regression Study

**DOI:** 10.64898/2026.09.08.26362487

**Authors:** Nhan T. Ho

## Abstract

**Background:** BMI and height are usually related through mean-based models, which can mask how this relationship varies across the height distribution and across age. Whether BMI relates to height differently at different quantiles, and whether this differs by sex, is not well studied in Southeast Asian children.

**Methods:** We analyzed longitudinal school health data from 32,006 Vietnamese children (16,489 boys, 15,517 girls) aged 5 to 18 with at least 3 annual visits from 2018 to 2025. Using sex-stratified linear quantile mixed models, we estimated the association between BMI-for-age Z-score (BMIZ) and height-for-age Z-score (HAZ) at 5 quantiles, adjusting for a flexible age spline and study site, with a child-level cluster bootstrap for inference, plus a pooled model with a formal sex interaction test.

**Results:** A 1SD increase in BMIZ was linked to higher HAZ at every quantile in both sexes (boys 0.078 to 0.100, girls 0.055 to 0.089, all p < .01), with no significant sex interaction (p = .35), contrasting with a small negative association from a linear mixed model (−0.023). This association did not differ significantly across quantiles within either sex, pointing to a fairly uniform effect across the height distribution itself, but it did vary strongly by age, strongest in early and mid-childhood and reversing to negative by late adolescence in both sexes, so children with overweight or obesity were taller for age through most of childhood but shorter than leaner peers by the oldest ages studied. Within-group variability in HAZ narrowed with age in both sexes, and boys with thinness showed a consistently wider height spread than boys in the other BMI categories through most of childhood, a pattern that was less pronounced in girls.

**Conclusions:** BMI’s association with linear growth in Vietnamese children is positive through most of childhood but weakens and reverses by late adolescence, a pattern mean-based models obscure. BMI and height should be interpreted together in light of a child’s age, not as one fixed relationship.

## Introduction

Height for age is one of the most widely used markers of child health, and body mass index is routinely treated as little more than a size correction when that marker is interpreted. Most published work still summarizes the BMI to height relationship as a single average effect, even though children near the short and tall ends of the height distribution may respond quite differently to being under or over their expected weight. A growing body of quantile regression work suggests this average based view hides real heterogeneity. Among Indian children under three, maternal education, birth order, and economic status shaped the lower tail of the height for age distribution differently than the upper tail ^1^. In British birth cohorts followed from 1953 to 2015, socioeconomic gaps in BMI were roughly two and a half times larger at the 90th percentile than at the 50^th 2^, and a comparable pattern was seen in China, where urban rural BMI differences vanished at the lowest quantile but persisted at the highest across three decades of surveillance ^3^.

Part of the reason BMI behaves unevenly across the height distribution is biological rather than statistical. A global analysis spanning 200 countries and 65 million participants found that height and BMI trajectories diverge substantially by country, sex, and age, pointing to systematic heterogeneity in how weight and height track together during growth ^4^. Earlier work from the same consortium showed that BMI trends in children have accelerated sharply across parts of Asia while plateauing in many high income countries ^5^. At the individual level, allometric analyses of several million children have shown that weight does not scale to height with a fixed exponent of two as BMI assumes, with the exponent drifting most in mid childhood before stabilizing near age 17 ^6^, and that standard underweight and overweight cutoffs misclassify shorter children several times more often than taller peers of the same age ^7^. BMI can also reflect either fat mass or fat free mass gain during puberty, further limiting its meaning as a single number ^8^. Japanese children who became obese by age 15 showed earlier peak height velocity than their peers, suggesting BMI trajectories and pubertal height timing are mechanistically linked rather than independent ^9^.

Vietnam illustrates why this matters for a population undergoing rapid nutritional transition. Among urban Vietnamese schoolchildren, the combined prevalence of overweight and obesity has reached roughly three times the WHO threshold for very high public health concern in boys, with clear regional differences in weight status and height across Hanoi, Ho Chi Minh City, and Haiphong ^10^. Rural to urban BMI gaps have widened over the past two decades ^11^, and maternal nutritional status before conception continues to shape offspring linear growth years later ^12^. Quantile regression has already been applied to height for age in India and to BMI in Britain and China, but to our knowledge no study has used it to examine how BMI differentially shapes the conditional height distribution in a Southeast Asian population, leaving a clear gap where growth patterns do not simply mirror international references.

We therefore used a large longitudinal school based cohort in Vietnam to characterize how BMI relates to the entire conditional distribution of height for age (not just its mean) across ages around puberty for boys and girls using quantile mixed models.

## Methods

We performed retrospective analysis of longitudinal anthropometric data from annual school health check from 2018 to 2025 of children aged 5 to 18 years attending a private school system in three major cities in Vietnam (Hanoi, Hochiminh and Haiphong). The study was approved by Vinmec Ethical Committee (approval number 0231/2024/CN/HDDD VMEC). Informed consent was waived for retrospective analysis of de-identified data.

Children were included if they had at least three annual visits recorded, giving each child enough repeated measurements to support a mixed model approach. Height for age Z scores (HAZ) and BMI for age Z scores (BMIZ) were calculated against the WHO 2007 growth reference for school aged children and adolescents ^13^. Records with implausible values, HAZ or BMIZ beyond plus or minus 6 SD, were excluded. BMIZ was also converted into four clinical categories, thinness, normal weight, overweight, and obesity, using standard WHO cut points.

Our primary analysis modeled the relationship between BMIZ and HAZ across the full conditional height distribution, not just its mean, using linear quantile mixed models fit with the lqmm package in R ^14,15^. Because boys and girls differ in the timing and tempo of pubertal growth, we fit these models separately for each sex rather than pooling them with sex as a simple covariate. Each model regressed HAZ on BMIZ interacted with a natural cubic spline of standardized age (4 degrees of freedom), adjusting for study site, with a random intercept for each child. Models were fit across a grid of quantiles from 0.05 to 0.95, with five headline quantiles (0.05, 0.25, 0.50, 0.75, 0.95) carried forward for inference. Ninety five percent confidence intervals (95%CI) and two sided p values came from a child level cluster bootstrap, resampling whole children rather than individual visits to respect the repeated measures structure ^16^. We tested whether the BMIZ effect varied meaningfully across quantiles with a Wald test built from the bootstrap covariance of the headline coefficients, and summarized the extremes of the distribution as the difference between the 95th and 5th percentile coefficients. An age by quantile heatmap and predicted HAZ curves by BMI category were generated from the fitted models using a custom prediction routine built to reconstruct the spline basis correctly for new data, since R’s standard prediction tools for this model class do not handle derived terms like splines reliably. Distributional inequality across BMI categories was summarized as the P95 minus P5 spread and the interquartile range of predicted HAZ at each age.

Secondary analyses included the same quantile model fit to the pooled cohort with sex added as a covariate, and a linear mixed model, both fit on the pooled cohort, to place the distributional findings alongside more conventional mean based approaches. We also formally tested for a sex by BMIZ interaction using a single pooled model that allowed both the BMIZ slope and the shape of the age spline to differ by sex, again with bootstrap based inference. As sensitivity analyses, we refit the headline quantiles with a quantile generalized additive model, qgam ^17^, and with a naive quantile regression that ignored within child clustering ^18^, separately for boys, girls, and the pooled cohort, and checked whether results were stable when the spline was given 3, 4, or 5 degrees of freedom instead of 4.

All analyses were run in R version 4.5.1 ^19^, with quantile grid fitting and bootstrap replicates parallelized using the furrr ^20^ and future ^21^ packages. Statistical significance was set at a two sided p value below 0.05.

## Results

The cohort included 32,006 children, 16,489 boys and 15,517 girls, each with a median of 4 annual visits (interquartile range 3 to 5) (**Table 1**). Boys and girls entered the cohort at a similar median age (7.6 and 7.8 years) and had a comparable mean HAZ (0.33 and 0.17), but boys had a substantially higher mean BMIZ at entry than girls (0.83 versus 0.21) and a much higher combined prevalence of overweight and obesity (47.1% versus 27.8%) (**Table 1**). Study site distribution was similar between sexes, with roughly two thirds of children from the Hanoi site, one quarter from Ho Chi Minh City, and the remainder from Haiphong (**Table 1**).

**Table 1.** Baseline characteristics of the cohort, by sex.

| Characteristic | Boys (n = 16,489) | Girls (n = 15,517) |
| --- | --- | --- |
| Visits per child, median (IQR) | 4 (3 to 5) | 4 (3 to 5) |
| Age at entry, years, median (IQR) | 7.6 (6.1 to 10.5) | 7.8 (6.1 to 10.8) |
| HAZ at entry, mean (SD) | 0.33 (0.97) | 0.17 (0.95) |
| BMIZ at entry, mean (SD) | 0.83 (1.49) | 0.21 (1.23) |
| Study site, n (%) |  |  |
| Hanoi | 10,869 (65.9) | 10,076 (64.9) |
| Ho Chi Minh City | 4,244 (25.7) | 4,221 (27.2) |
| Haiphong | 1,376 (8.3) | 1,220 (7.9) |
| **BMI category at entry, n (%)** |  |  |
| Thinness | 405 (2.5) | 512 (3.3) |
| Normal BMI | 8,301 (50.4) | 10,670 (68.9) |
| Overweight | 3,730 (22.6) | 3,179 (20.5) |
| Obesity | 4,036 (24.5) | 1,131 (7.3) |

Values are median (interquartile range), mean (SD), or n (%), measured at each child's first recorded visit. HAZ= height-for-age Z-score, BMIZ= BMI-for-age Z-score, IQR= interquartile range, SD= standard deviation. BMI categories were defined from BMIZ using WHO cutoffs: thinness BMIZ < -2SD, normal BMI -2 to < 1, overweight 1 to < 2, obesity $\geq 2$ .

The pooled linear mixed model, which summarizes the BMIZ-HAZ relationship as one mean effect adjusted linearly for age, found a small negative association, a 0.023 SD lower HAZ per 1-SD increase in BMIZ (95% CI −0.027 to −0.019) (**Table 2**). This was the opposite direction from every quantile-based model in this study. The pooled linear quantile mixed model, which adjusts for age through a flexible spline rather than a simple linear term, found a positive association at all five headline quantiles, ranging from 0.043 to 0.075 (**Table 2**, **Figure 1**).

**Figure 1.**
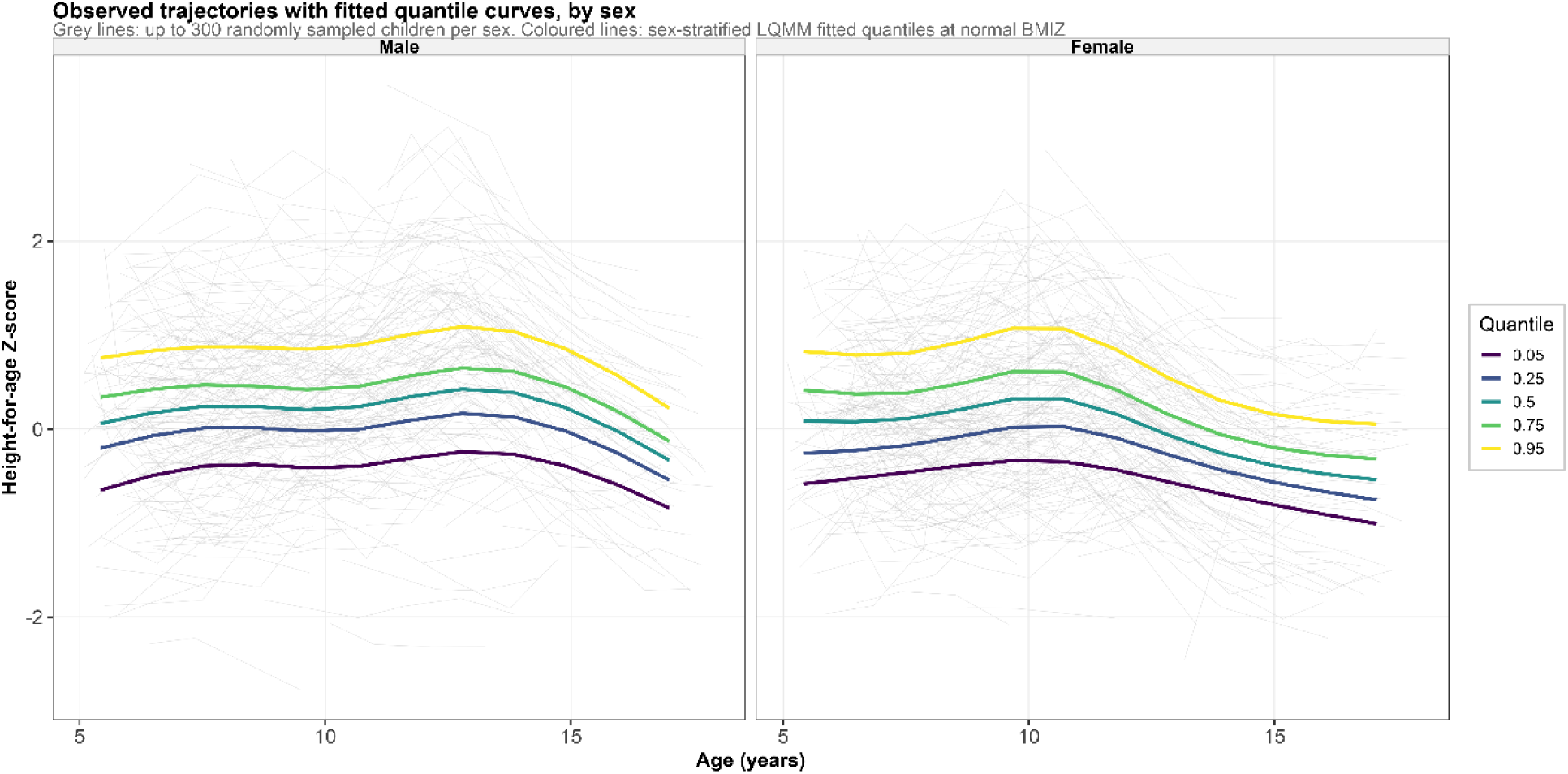
Observed height-for-age trajectories with fitted quantile curves, by sex. Grey lines show the raw longitudinal HAZ trajectories of up to 300 randomly sampled children per sex. Colored lines show the fitted 5th, 25th, 50th, 75th, and 95th percentile curves from the sex-stratified linear quantile mixed models (Table 3), predicted at a normal BMIZ (0) for a representative child at the Hanoi site.

**Table 2.**
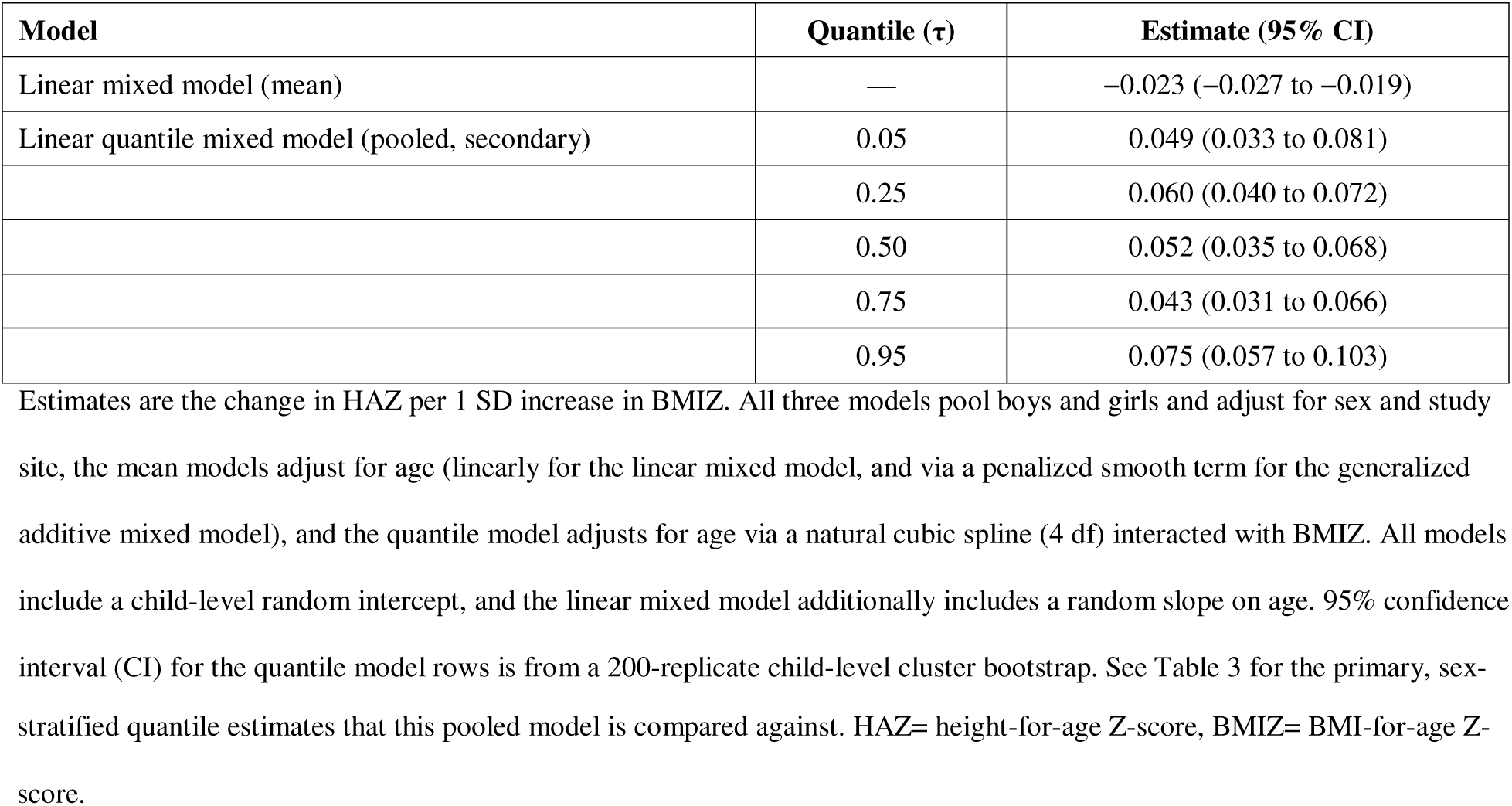
BMIZ-HAZ association in the pooled cohort: mean-based models versus the distributional (quantile) model.

| Model | Quantile ($\tau$) | Estimate (95% CI) |
| --- | --- | --- |
| Linear mixed model (mean) | — | -0.023 (-0.027 to -0.019) |
| Linear quantile mixed model (pooled, secondary) | 0.05 | 0.049 (0.033 to 0.081) |
|  | 0.25 | 0.060 (0.040 to 0.072) |
|  | 0.50 | 0.052 (0.035 to 0.068) |
|  | 0.75 | 0.043 (0.031 to 0.066) |
|  | 0.95 | 0.075 (0.057 to 0.103) |

stratified quantile estimates that this pooled model is compared against. HAZ= height-for-age Z-score, BMIZ= BMI-for-age Z-score.

**Table 3.** Association between BMIZ and HAZ across the conditional height-for-age distribution: primary (sex-stratified) and secondary (pooled) linear quantile mixed models.

| Sex / model | Quantile ($\tau$) | Estimate (95% CI) | p<sup>□</sup> |
| --- | --- | --- | --- |
| Boys (primary) | 0.05 | 0.081 (0.041 to 0.101) | < .01 |
|  | 0.25 | 0.089 (0.062 to 0.105) | < .01 |
|  | 0.50 | 0.078 (0.059 to 0.104) | < .01 |
|  | 0.75 | 0.083 (0.068 to 0.120) | < .01 |
|  | 0.95 | 0.100 (0.073 to 0.143) | < .01 |
|  | Difference between the 95th- and 5th-percentile coefficients | 0.041 (−0.000 to 0.080) |  |
| Girls (primary) | 0.05 | 0.079 (0.051 to 0.136) | < .01 |
|  | 0.25 | 0.055 (0.030 to 0.077) | < .01 |
|  | 0.50 | 0.066 (0.043 to 0.090) | < .01 |
|  | 0.75 | 0.061 (0.030 to 0.097) | < .01 |
|  | 0.95 | 0.089 (0.049 to 0.114) | < .01 |
|  | Difference between the 95th- and 5th-percentile coefficients | −0.004 (−0.054 to 0.043) |  |
| Pooled (secondary) | 0.05 | 0.049 (0.033 to 0.081) | < .01 |
|  | 0.25 | 0.060 (0.040 to 0.072) | < .01 |
|  | 0.50 | 0.052 (0.035 to 0.068) | < .01 |
|  | 0.75 | 0.043 (0.031 to 0.066) | < .01 |
|  | 0.95 | 0.075 (0.057 to 0.103) | < .01 |
|  | Difference between the 95th- and 5th-percentile coefficients | 0.024 (−0.003 to 0.057) |  |

Estimates are the change in HAZ per 1-SD increase in BMIZ, from linear quantile mixed models that adjust for a natural cubic spline of age (4 df) interacted with BMIZ, study site, and a child-level random intercept. The primary models are fit separately for boys and girls; the secondary pooled model additionally adjusts for sex. 95% CI and p values are from a 200-replicate child-level cluster bootstrap. □ Two-sided bootstrap p value; reported as < .01 where none of the 200 bootstrap replicates crossed zero. Quantile-heterogeneity Wald test, testing whether the BMIZ effect differs across the 5 quantiles within a stratum: boys $\chi^2 = 1.69$ , df = 4, p = .79; girls $\chi^2 = 4.39$ , df = 4, p = .36; pooled $\chi^2 = 6.41$ , df = 4, p = .17. Difference between the 95th- and 5th-percentile coefficients: boys 0.041 (−0.000 to 0.080); girls −0.004 (−0.054 to 0.043); pooled 0.024 (−0.003 to 0.057).

In the primary, sex-stratified analysis, a 1 SD increase in BMIZ was associated with higher HAZ at every headline quantile in both sexes, and every one of these ten estimates was significant at p < .01 (**Table 3**). The association was somewhat larger and more consistent in boys, ranging from 0.078 to 0.100 across the five quantiles, than in girls, where it ranged from 0.055 to 0.089 (**Table 3**, **Figure 2A**). Within each sex, the BMIZ effect did not differ significantly across quantiles (quantile-heterogeneity Wald test for boys p = .79, for girls p = .36), and the same held for the pooled secondary model (p = .17) (**Table 3**). The difference between the 95th- and 5th-percentile coefficients crossed zero (not statistically significant) in every stratum (boys 0.041, girls −0.004, pooled 0.024) (**Table 3**), so we found no strong evidence that BMI’s association with height differs between short and tall children once age is properly accounted for.

**Figure 2.**
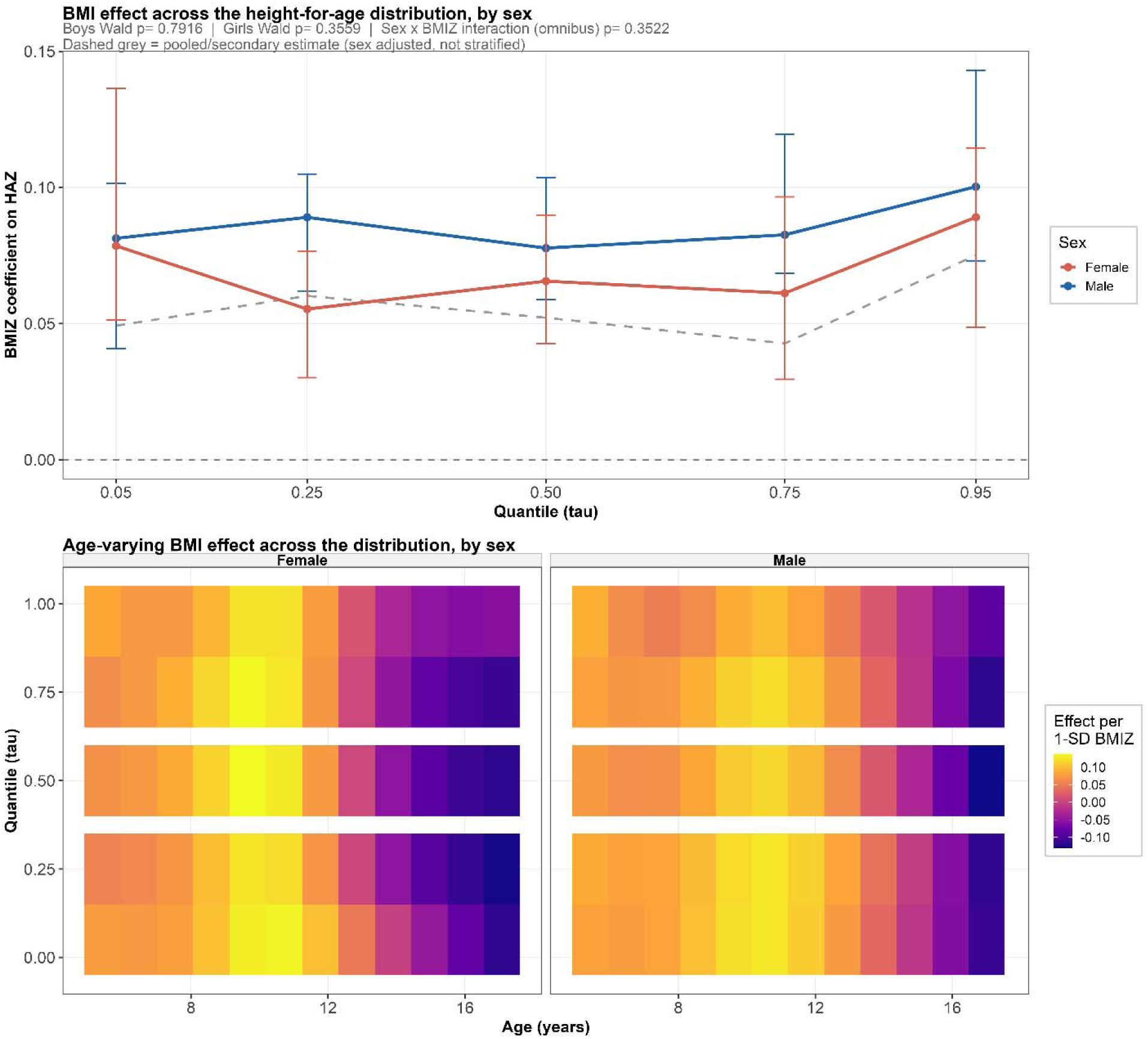
BMI effect across the height-for-age distribution and across age, by sex. (A) BMIZ coefficient (change in HAZ per 1-SD increase in BMIZ) at each of the 5 headline quantiles for boys and girls, with 95% bootstrap CI (Table 3). The dashed grey line is the pooled/secondary estimate, not stratified by sex, for comparison (Table 2). The Wald p values test whether the BMIZ effect varies across quantiles within each sex; the sex × BMIZ interaction p value is the omnibus test reported in Table 4. (B) Heatmap of the model-implied change in HAZ for a 1-SD increase in BMIZ across age and quantile, separately by sex. Yellow indicates a positive association between BMI and height-for-age; purple indicates a negative association.

**Table 4.**
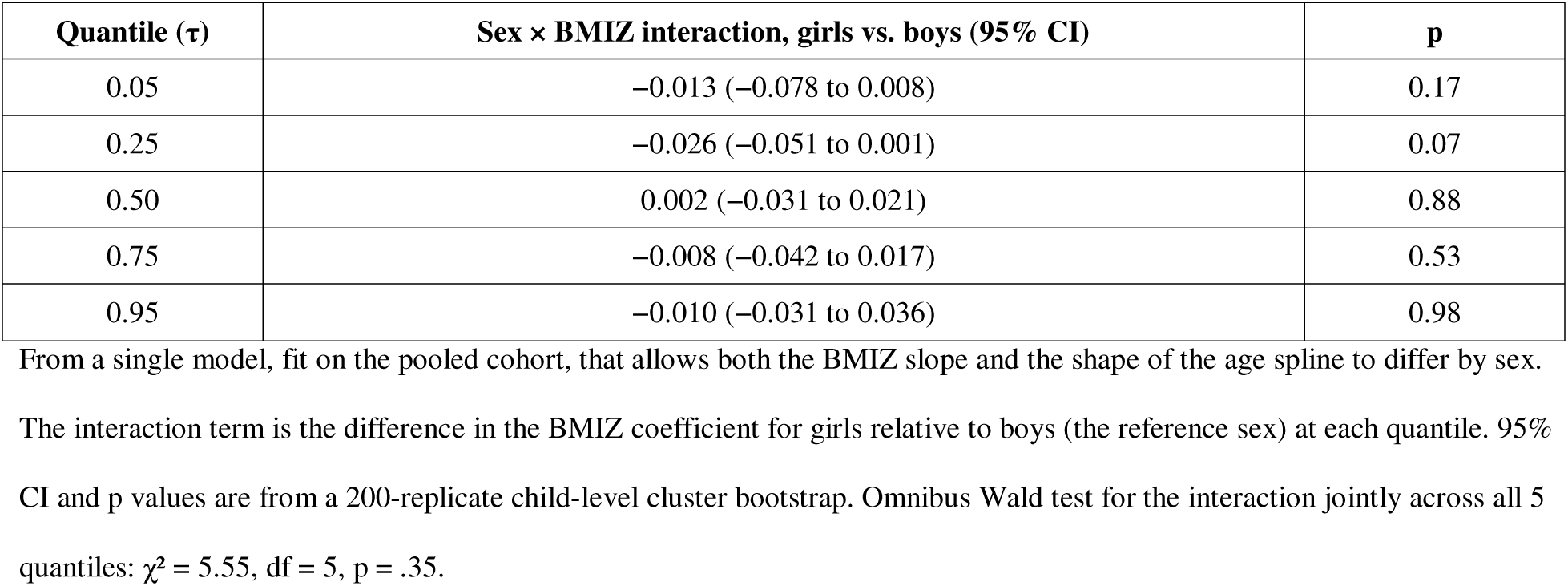
Formal test for a sex × BMIZ interaction in the association with HAZ.

A formal test for a sex by BMIZ interaction, fit in a single pooled model that let both the BMIZ slope and the shape of the age spline differ by sex, found the interaction estimate was negative at four of the five quantiles, meaning the BMIZ effect was estimated slightly lower in girls than boys everywhere except the median, but none of the five per-quantile tests reached significance, the closest being tau = 0.25 (estimate −0.026, 95% CI −0.051 to 0.001, p = .07) (**Table 4**). The omnibus Wald test for the interaction jointly across all five quantiles was also not significant (p = .35) (**Table 4**). So while boys showed numerically higher point estimates throughout **Table 3**, this sex difference did not reach statistical significance in our formal test.

The age by quantile heatmap (**Figure 2B**) showed that the positive BMIZ-HAZ association in **Table 3** was not stable across childhood. In both sexes, higher BMIZ was linked to greater height-for-age in early and mid-childhood, roughly ages 6 to 12, but this association weakened with age and reversed to negative by around age 15 to 17. This same reversal appeared directly in the predicted growth curves (**Figure 3**). At every headline quantile, children with overweight or obesity were predicted to be taller for their age than children with thinness through most of childhood, but the curves converged in mid-adolescence and then crossed, so that by the oldest ages studied children with thinness or normal BMI were taller for age than children with obesity.

**Figure 3.**
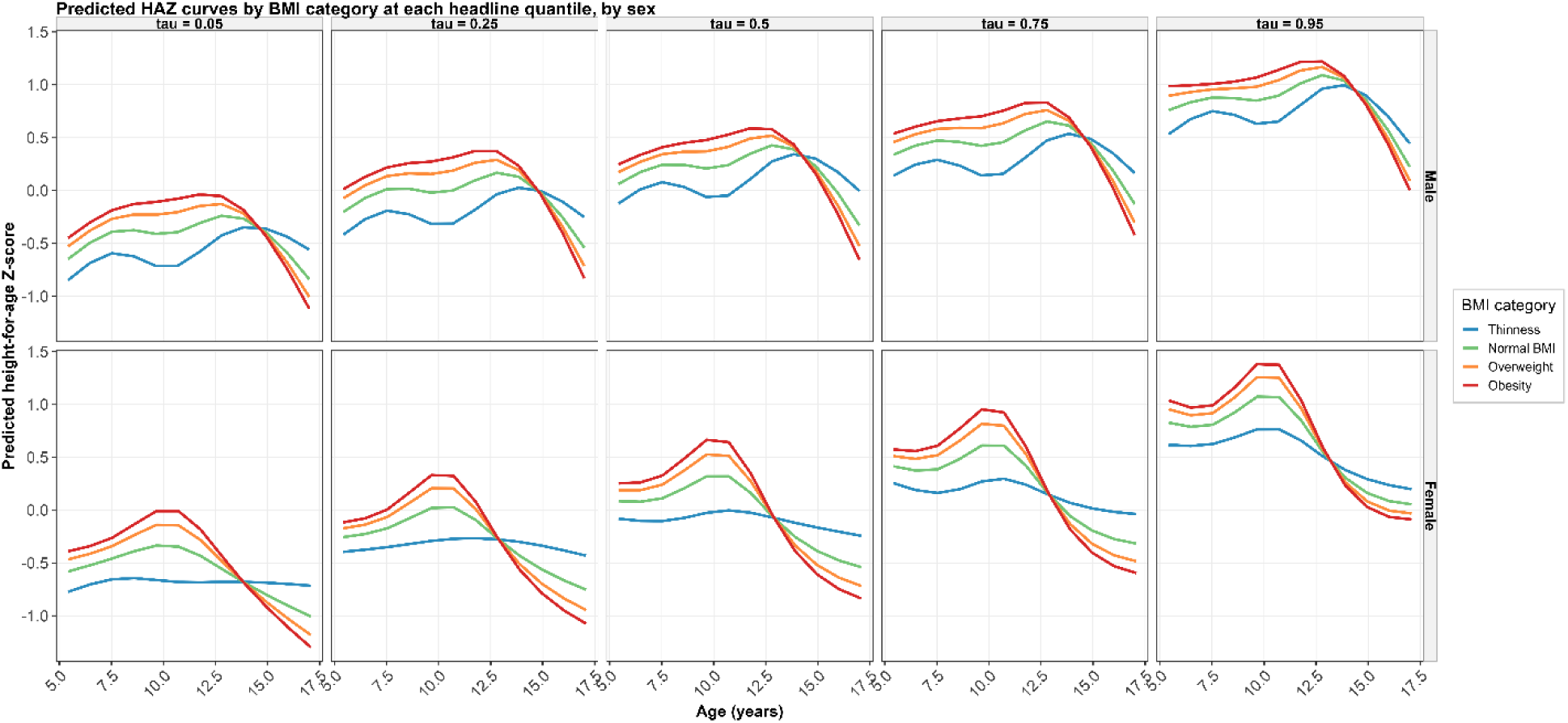
Predicted height-for-age curves by BMI category, at each headline quantile, by sex. Each panel shows the predicted HAZ trajectory for a representative child in each of four BMI categories (thinness, normal BMI, overweight, obesity, represented by BMIZ values of −2.5, 0, 1.5, and 2.5 respectively), at a given quantile (columns) and sex (rows), holding study site (Hanoi) fixed.

Within-group variability in HAZ (**Figure 4**) generally narrowed with age in both sexes, consistent with the pubertal convergence of height seen in **Figure 3**, though it widened again modestly at the oldest ages, most visibly in girls with obesity. In boys, children with thinness showed a consistently wider HAZ spread, both the P95-P5 range and the interquartile range, than the other three BMI categories through most of childhood, a pattern that was much less pronounced in girls, where the four BMI categories tracked closely together until adolescence.

**Figure 4.**
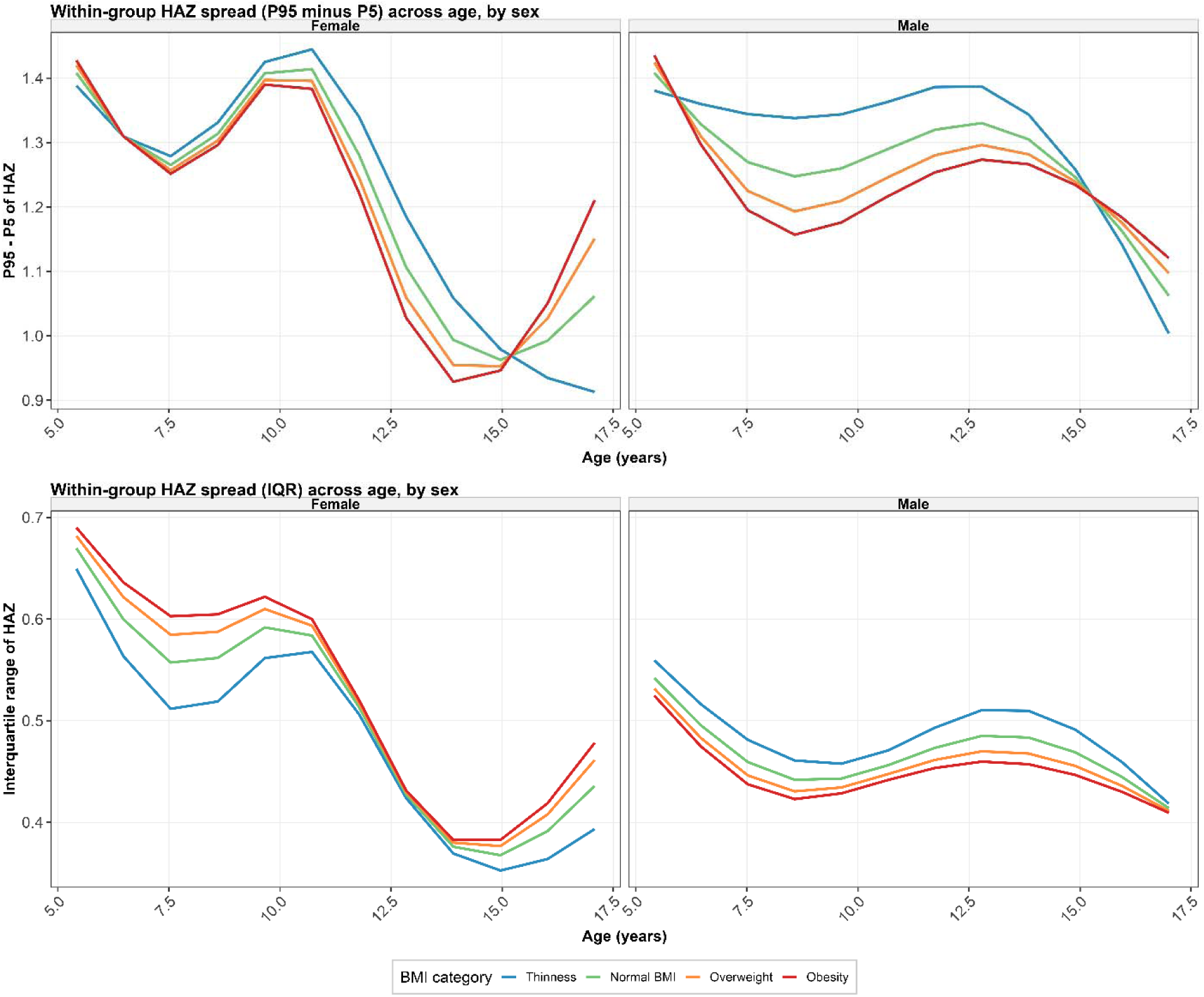
Within-group spread of the height-for-age distribution across age, by BMI category and sex. Top row: difference between the 95th and 5th percentile of predicted HAZ (P95 − P5) across age, by BMI category and sex. Bottom row: interquartile range (P75 − P25) of predicted HAZ, shown the same way.

Model diagnostics supported the primary analysis. No quantile crossing was observed in any stratum, and residuals from the median models were reasonably well behaved, though with somewhat heavier tails than a normal distribution (**Figure S1**). Six of the 15 quantile-by-stratum primary models did not fully meet the convergence criterion even after a second fitting attempt with a higher iteration limit, four of these six at the most extreme quantiles, tau = 0.05 or 0.95, where data are sparser, and the remaining two at tau = 0.75 in boys and tau = 0.05 in girls and the pooled model (**Table S1**). None of the 200 bootstrap replicates failed to converge at any quantile in any stratum, so the confidence intervals in **Table 3** are based on the full bootstrap sample throughout, with bootstrap standard errors ranging from about 0.009 to 0.023 (**Table S2**).

Sensitivity analyses supported the direction, though not the exact magnitude, of the primary findings. A quantile generalized additive model and a naive quantile regression that ignored within-child clustering both agreed with the primary LQMM that the BMIZ-HAZ association was positive at every quantile and in every stratum, but both produced point estimates roughly three to four times larger, in the range of 0.17 to 0.28 compared with 0.04 to 0.10 for the primary model (**Figure S1, Table S3**), consistent with the expected effect of ignoring repeated-measures clustering on point estimates as well as precision. The median-quantile BMIZ coefficient was directionally stable but showed some sensitivity to how flexible the age spline was allowed to be, higher with 4 degrees of freedom, the primary specification, than with 3 or 5 in both boys (0.078 at 4 df versus 0.054 at 3 df and 0.069 at 5 df) and girls (0.066 at 4 df versus 0.031 at 3 df and 0.060 at 5 df) (**Table S4**).

## Discussion

This large, sex-stratified analysis of Vietnamese schoolchildren found that a higher BMI-for-age was associated with a taller height-for-age across the entire conditional height distribution, not just on average, and that this association changed direction with age rather than staying constant across childhood and adolescence. The positive association was strongest in early and mid-childhood and reversed to a negative one by mid-to-late adolescence, a pattern nearly identical in boys and girls even though the formal interaction test did not reach statistical significance.

The contrast between our pooled linear mixed model, which found a small negative average association, and our quantile models, which found a consistently positive one once age was modeled flexibly, illustrates why relying on a single mean estimate can be misleading in growth research. Quantile regression has previously revealed determinants of child growth that a mean model would have missed, for maternal education and birth order in Indian toddlers ^1^ and for maternal age and birth weight in Ethiopian preschoolers ^22^. Our results extend this logic in a different direction. Rather than the BMI-height association varying across the height distribution itself, which our quantile heterogeneity tests found little evidence for within either sex, the more important source of heterogeneity in this cohort was age. A model that assumes one linear BMI-age interaction for the whole growth period will average over a relationship that is actually reversing sign, which likely explains the discrepancy with the linear mixed model.

The age-varying reversal we observed is biologically plausible given what is known about BMI and pubertal timing. Japanese adolescents who became obese by age 15 showed earlier adiposity rebound and a steeper rise in BMI through puberty than their peers ^9^, and earlier pubertal onset is generally accompanied by an earlier, shorter growth spurt and earlier epiphyseal closure. If higher-BMI children in our cohort matured earlier, they may have been taller for their age in childhood but then plateaued sooner, while later-maturing, lower-BMI peers kept growing into late adolescence and closed the gap. This interpretation fits evidence that BMI’s relationship with body composition is itself age-dependent, since the ratio of weight to height squared does not scale consistently across childhood and adolescence and BMI-based weight categories misclassify children differently depending on height ^6,7^, and BMI cannot separate fat mass from lean mass at any single age ^8^, so the same BMIZ value likely reflects different combinations of fat, muscle, and skeletal maturity at age 6 than at age 16.

The higher and more consistent BMI-height association we found in boys, alongside their much higher combined prevalence of overweight and obesity than girls, fits a broader regional pattern. Our group’s earlier analysis of this same school system found high overweight and obesity prevalence with meaningful differences across Hanoi, Ho Chi Minh City, and Haiphong ^10^, and BMI trends in children have been rising faster in parts of Asia than in most high income countries even as those countries’ own trends have leveled off ^5^. Height and BMI trajectories vary substantially by country ^4^ (9), and China’s urban-rural gap in child height and BMI has been narrowing as both groups rise together ^3^, which is a reason to be cautious about generalizing our findings beyond the private, urban school population studied here.

These findings matter beyond growth monitoring itself. Higher BMI in youth predicts higher adult coronary heart disease risk while greater height predicts lower risk ^23^, so a period in which BMI and height move together in the same direction, followed by a period in which they diverge, could carry different long-term implications depending on when it is measured. Clinicians and researchers interpreting a child’s height-for-age alongside their BMI should expect the relationship between the two to depend heavily on the child’s age.

This study has several limitations. A meaningful minority of quantile models, concentrated mostly at the extreme tails, did not fully meet the convergence criterion even after a second fitting attempt, and those estimates should be interpreted cautiously. The cohort spans 2018 to 2025, a period that includes documented pandemic-era shifts in child BMI ^24^, which we could not separate from other secular trends. Finally, as an observational cohort, we cannot rule out that faster-maturing children drive both their own BMI and their own height trajectory, rather than BMI itself shaping height. Our companion work extending this distributional approach in this cohort (which is presented in a separate paper) incorporates direct measures of pubertal timing and tempo, such as age at peak height velocity, to test whether earlier maturation explains the age-varying BMI-height association reported here.

Taken together, these findings suggest that a single overweight or obesity label carries a different meaning for a child’s height depending on age. In routine growth monitoring, a young child who is both heavy and tall for age may simply be on a faster-maturing trajectory rather than one that predicts a lasting height advantage, and clinicians should be cautious about reassuring families that a heavier child’s height edge over peers will persist into adolescence. From a public health standpoint, school-based growth surveillance programs like this one could add real value by tracking BMI and height together across childhood rather than at a single time point, since the crossover we observed would be invisible in a cross-sectional snapshot.

## Declaration

### Contributors’ statement

Nhan Thi Ho did conceptualization, data curation, formal analysis, investigation, methodology, project administration, resources, software, supervision, validation, visualization, writing original draft, and writing review & editing.

### Data Sharing Statement

R codes are available from the corresponding author upon reasonable request. Individual patient-level data cannot be shared due to applicable privacy regulations and the terms of the institutional ethics approval.

### Conflict of intertest

The sole author states that there is no conflict of interest.

### Ethic and Consent statement

This study was approved by Vinmec Ethical Committee (approval number 0231/2024/CN/HDDD VMEC) with a waiver of individual informed consent as the study used de-identified routinely collected retrospective school health check data.

### Use of Artificial Intelligence

The authors performed all original research work regarding scientific content, analyses, interpretations and manuscript writing. The authors used AI-assisted tools for language editing and grammar checking during manuscript preparation.

## References

1. Mokalla, T. R. & Mendu, V. V. R. Application of quantile regression to examine changes in the distribution of Height for Age (HAZ) of Indian children aged 0–36 months using four rounds of NFHS data. PLoS One 17, (2022).

2. Bann, D., Johnson, W., Li, L., Kuh, D. & Hardy, R. Socioeconomic inequalities in childhood and adolescent body-mass index, weight, and height from 1953 to 2015: an analysis of four longitudinal, observational, British birth cohort studies. Lancet Public Health 3, (2018).

3. Luo, D. et al. Long-term trends and urban-rural disparities in the physical growth of children and adolescents in China: an analysis of five national school surveys over three decades. Lancet Child Adolesc. Health (2023) doi:10.1016/s2352-4642(23)00175-x.

4. Rodriguez-Martinez, A. et al. Height and body-mass index trajectories of school-aged children and adolescents from 1985 to 2019 in 200 countries and territories: a pooled analysis of 2181 population-based studies with 65 million participants. The Lancet 396, (2020).

5. Abarca-Gómez, L. et al. Worldwide trends in body-mass index, underweight, overweight, and obesity from 1975 to 2016: a pooled analysis of 2416 population-based measurement studies in 128·9 million children, adolescents, and adults. The Lancet 390, 2627–2642 (2017).

6. Ogata, H. et al. Allometric multi-scaling of weight-for-height relation in children and adolescents: Revisiting the theoretical basis of body mass index of thinness and obesity assessment. PLoS One 19, (2024).

7. Isoyama, Y. et al. Age- and height-dependent bias of underweight and overweight assessment standards for children and adolescents. Front. Public Health 12, (2024).

8. Chung, S. Body mass index and body composition scaling to height in children and adolescent. Ann. Pediatr. Endocrinol. Metab. 20, 125–129 (2015).

9. Matsumoto, N. et al. Trajectory of body mass index and height changes from childhood to adolescence: a nationwide birth cohort in Japan. Sci. Rep. 11, (2021).

10. Ho, N. T. et al. Overweight & obesity epidemic, temporal trends and regional disparities in physical growth of Vietnamese children. Sci. Rep. 16, 7515 (2026).

11. Wake, S., Zewotir, T., Mekebo, G. & Fissuh, Y. H. Rural-urban differentials in child body mass index over time. BMC Pediatr. 23, (2023).

12. Young, M. et al. Role of maternal preconception nutrition on offspring growth and risk of stunting across the first 1000 days in Vietnam: A prospective cohort study. PLoS One 13, (2018).

13. De Onis, M. et al. Development of a WHO growth reference for school-aged children and adolescents. Bull. World Health Organ. 85, (2007).

14. Geraci, M. & Bottai, M. Linear quantile mixed models. Stat. Comput. 24, (2014).

15. Geraci, M. Linear quantile mixed models: The lqmm package for laplace quantile regression. J. Stat. Softw. 57, (2014).

16. Colin Cameron, A., Gelbach, J. B. & Miller, D. L. Bootstrap-based improvements for inference with clustered errors. Review of Economics and Statistics 90, (2008).

17. Fasiolo, M., Wood, S. N., Zaffran, M., Nedellec, R. & Goude, Y. qgam: Bayesian Nonparametric Quantile Regression Modeling in R. J. Stat. Softw. 100, (2021).

18. Koenker, R. & Bassett, G. Regression Quantiles. Econometrica 46, (1978).

19. R Core Team. R: A Language and Environment for Statistical Computing. R Foundation for Statistical Computing, Vienna, Austria Preprint at 10.1017/CBO9781107415324.004 (2016).

20. Vaughan, D., Bengtsson, H. & Dancho, M. furrr: Apply Mapping Functions in Parallel using Futures. Preprint at 10.32614/CRAN.package.furrr (2018).

21. Bengtsson, H. A Unifying Framework for Parallel and Distributed Processing in R using Futures. R Journal 13, (2021).

22. Yirga, A., Ayele, D. & Melesse, S. Application of Quantile Regression: Modeling Body Mass Index in Ethiopia. Open Public Health J. (2018) doi:10.2174/1874944501811010221.

23. Meyer, J. et al. Associations between body mass index and height during childhood and adolescence and the risk of coronary heart disease in adulthood: A systematic review and meta analysis. Obesity Reviews 22, (2021).

24. Woolford, S. et al. Changes in Body Mass Index Among Children and Adolescents During the COVID-19 Pandemic. JAMA (2021) doi:10.1001/jama.2021.15036.

