## Supplementary Tables and Figures for "The Age-Varying Association Between BMI and Height-for-Age in Vietnamese Schoolchildren: A Sex-Stratified Quantile Regression Study"

Nhan Thi Ho

**Table S1. Convergence status of linear quantile mixed models across the evaluated quantiles by analytical stratum.**

| **Stratum** | **Model** | **Converged** |
| --- | --- | --- |
| Boys (primary) | tau_0.05 | TRUE |
| Boys (primary) | tau_0.25 | TRUE |
| Boys (primary) | tau_0.50 | TRUE |
| Boys (primary) | tau_0.75 | FALSE |
| Boys (primary) | tau_0.95 | FALSE |
| Girls (primary) | tau_0.05 | FALSE |
| Girls (primary) | tau_0.25 | TRUE |
| Girls (primary) | tau_0.50 | TRUE |
| Girls (primary) | tau_0.75 | TRUE |
| Girls (primary) | tau_0.95 | FALSE |
| Pooled (secondary) | tau_0.05 | FALSE |
| Pooled (secondary) | tau_0.25 | TRUE |
| Pooled (secondary) | tau_0.50 | TRUE |
| Pooled (secondary) | tau_0.75 | TRUE |
| Pooled (secondary) | tau_0.95 | FALSE |

This table summarizes whether the models met the convergence criteria across the full quantile grid for both the primary sex-stratified models and the secondary pooled cohort model. Estimation algorithms included a second fitting attempt with a higher iteration limit for initial non-convergences. Four of the six ultimate convergence failures occurred at the extreme 0.05 and 0.95 quantiles where data are sparser.

**Table S2. Bootstrap stability and standard errors at headline quantiles for each stratum.**

| **Stratum** | **Tau (quantile)** | **Estimate** | **Bootstrap lower 95%CI** | **Bootstrap upper 95%CI** | **Bootstrap SE** | **Number of valid replicates** | **Number failed** | **Number success** |
| --- | --- | --- | --- | --- | --- | --- | --- | --- |
| Boys (primary) | 0.05 | 0.08131639 | 0.04080742 | 0.10143443 | 0.01527808 | 200 | 0 | 200 |
| Boys (primary) | 0.25 | 0.08903072 | 0.06196273 | 0.10492728 | 0.0108888 | 200 | 0 | 200 |
| Boys (primary) | 0.5 | 0.07772696 | 0.05887437 | 0.10365459 | 0.0114201 | 200 | 0 | 200 |
| Boys (primary) | 0.75 | 0.08260303 | 0.06843991 | 0.11950357 | 0.01353262 | 200 | 0 | 200 |
| Boys (primary) | 0.95 | 0.10027404 | 0.07307485 | 0.1429596 | 0.01749713 | 200 | 0 | 200 |
| Girls (primary) | 0.05 | 0.07854846 | 0.05131733 | 0.13644503 | 0.02258333 | 200 | 0 | 200 |
| Girls (primary) | 0.25 | 0.0553315 | 0.03013744 | 0.0766474 | 0.01333043 | 200 | 0 | 200 |
| Girls (primary) | 0.5 | 0.06558158 | 0.04268489 | 0.08989018 | 0.01265688 | 200 | 0 | 200 |
| Girls (primary) | 0.75 | 0.06117248 | 0.02953904 | 0.09655434 | 0.01690656 | 200 | 0 | 200 |
| Girls (primary) | 0.95 | 0.08905973 | 0.0485507 | 0.11449863 | 0.01698644 | 200 | 0 | 200 |
| Pooled (secondary) | 0.05 | 0.04921575 | 0.03298252 | 0.08105634 | 0.01280294 | 200 | 0 | 200 |
| Pooled (secondary) | 0.25 | 0.06019706 | 0.03991467 | 0.07193723 | 0.00868883 | 200 | 0 | 200 |
| Pooled (secondary) | 0.5 | 0.05216643 | 0.03495424 | 0.06753966 | 0.00832923 | 200 | 0 | 200 |
| Pooled (secondary) | 0.75 | 0.04271548 | 0.03069679 | 0.06594976 | 0.00997612 | 200 | 0 | 200 |
| Pooled (secondary) | 0.95 | 0.07512399 | 0.05723195 | 0.10286457 | 0.01112167 | 200 | 0 | 200 |

This table presents the standard errors and 95 percent confidence intervals at the five headline quantiles. Values were derived from a 200 replicate cluster bootstrap that resampled whole children to correctly account for the repeated measures data structure. All 200 bootstrap replicates successfully converged across all evaluated strata and quantiles. CI= confidence interval, SE= standard error.

**Table S3. Sensitivity analysis of the primary results compared against quantile generalized additive models and naive quantile regressions.**

| **Tau (quantile)** | **Method** | **Estimate** | **Lower 95%CI** | **Upper 95%CI** | **Stratum** |
| --- | --- | --- | --- | --- | --- |
| 0.05 | LQMM (primary) | 0.08131639 | 0.04080742 | 0.10143443 | Boys |
| 0.25 | LQMM (primary) | 0.08903072 | 0.06196273 | 0.10492728 | Boys |
| 0.5 | LQMM (primary) | 0.07772696 | 0.05887437 | 0.10365459 | Boys |
| 0.75 | LQMM (primary) | 0.08260303 | 0.06843991 | 0.11950357 | Boys |
| 0.95 | LQMM (primary) | 0.10027404 | 0.07307485 | 0.1429596 | Boys |
| 0.05 | LQMM (primary) | 0.07854846 | 0.05131733 | 0.13644503 | Girls |
| 0.25 | LQMM (primary) | 0.0553315 | 0.03013744 | 0.0766474 | Girls |
| 0.5 | LQMM (primary) | 0.06558158 | 0.04268489 | 0.08989018 | Girls |
| 0.75 | LQMM (primary) | 0.06117248 | 0.02953904 | 0.09655434 | Girls |
| 0.95 | LQMM (primary) | 0.08905973 | 0.0485507 | 0.11449863 | Girls |
| 0.05 | LQMM (secondary) | 0.04921575 | 0.03298252 | 0.08105634 | Pooled |
| 0.25 | LQMM (secondary) | 0.06019706 | 0.03991467 | 0.07193723 | Pooled |
| 0.5 | LQMM (secondary) | 0.05216643 | 0.03495424 | 0.06753966 | Pooled |
| 0.75 | LQMM (secondary) | 0.04271548 | 0.03069679 | 0.06594976 | Pooled |
| 0.95 | LQMM (secondary) | 0.07512399 | 0.05723195 | 0.10286457 | Pooled |
| 0.05 | qgam | 0.25360454 | 0.22982475 | 0.27738433 | Boys |
| 0.25 | qgam | 0.26233454 | 0.24446564 | 0.28020343 | Boys |
| 0.5 | qgam | 0.26329015 | 0.24790013 | 0.27868017 | Boys |
| 0.75 | qgam | 0.2775701 | 0.25903969 | 0.29610051 | Boys |
| 0.95 | qgam | 0.25685249 | 0.2273894 | 0.28631557 | Boys |
| 0.05 | rq (naive, ignores clustering) | 0.20145994 | 0.15112379 | 0.25179609 | Boys |
| 0.25 | rq (naive, ignores clustering) | 0.23870795 | 0.20860762 | 0.26880829 | Boys |
| 0.5 | rq (naive, ignores clustering) | 0.24609835 | 0.222799 | 0.2693977 | Boys |
| 0.75 | rq (naive, ignores clustering) | 0.23411922 | 0.20342538 | 0.26481305 | Boys |
| 0.95 | rq (naive, ignores clustering) | 0.22021509 | 0.16237701 | 0.27805317 | Boys |
| 0.05 | qgam | 0.2564276 | 0.23144312 | 0.28141207 | Girls |
| 0.25 | qgam | 0.26542998 | 0.24847714 | 0.28238281 | Girls |
| 0.5 | qgam | 0.24924648 | 0.2341911 | 0.26430185 | Girls |
| 0.75 | qgam | 0.23544167 | 0.21754748 | 0.25333587 | Girls |
| 0.95 | qgam | 0.20720489 | 0.18108777 | 0.23332202 | Girls |
| 0.05 | rq (naive, ignores clustering) | 0.27612735 | 0.20635503 | 0.34589966 | Girls |
| 0.25 | rq (naive, ignores clustering) | 0.22569966 | 0.19422966 | 0.25716967 | Girls |
| 0.5 | rq (naive, ignores clustering) | 0.22507257 | 0.18568065 | 0.26446449 | Girls |
| 0.75 | rq (naive, ignores clustering) | 0.22060671 | 0.17916477 | 0.26204865 | Girls |
| 0.95 | rq (naive, ignores clustering) | 0.17023658 | 0.11508316 | 0.22539001 | Girls |
| 0.05 | qgam | 0.24045971 | 0.22210517 | 0.25881425 | Pooled |
| 0.25 | qgam | 0.24015433 | 0.22741206 | 0.2528966 | Pooled |
| 0.5 | qgam | 0.23076481 | 0.21934218 | 0.24218745 | Pooled |
| 0.75 | qgam | 0.22772775 | 0.21391867 | 0.24153682 | Pooled |
| 0.95 | qgam | 0.21753301 | 0.19499573 | 0.24007029 | Pooled |
| 0.05 | rq (naive, ignores clustering) | 0.23007347 | 0.19832598 | 0.26182096 | Pooled |
| 0.25 | rq (naive, ignores clustering) | 0.22723186 | 0.20724377 | 0.24721996 | Pooled |
| 0.5 | rq (naive, ignores clustering) | 0.21789655 | 0.20147684 | 0.23431625 | Pooled |
| 0.75 | rq (naive, ignores clustering) | 0.21234636 | 0.19236194 | 0.23233078 | Pooled |
| 0.95 | rq (naive, ignores clustering) | 0.1920496 | 0.15405123 | 0.23004797 | Pooled |

This table compares the body mass index for age Z-score coefficients estimated by the primary models against two alternative modeling techniques. The comparison models include a quantile generalized additive model (qgam) and a naive quantile regression (rq). Both alternative methods ignored within-child clustering and subsequently produced point estimates approximately three to four times larger than the primary clustered approach. LQMM= Linear quantile mixed model (primary model separately for boys and girls, secondary model for pooled boys and girls). CI= confidence interval.

**Table S4. Sensitivity of the median regression coefficient to the degrees of freedom specified for the age spline.**

| **Stratum** | **Spline degree freedom** | **Estimate** |
| --- | --- | --- |
| Boys | 3 | 0.05415806 |
| Boys | 4 | 0.07772696 |
| Boys | 5 | 0.06857604 |
| Girls | 3 | 0.03103507 |
| Girls | 4 | 0.06613481 |
| Girls | 5 | 0.05981162 |

This table evaluates how the estimated body mass index coefficient changes when the natural cubic spline for standardized age is granted different levels of flexibility. The median quantile model was refit separately for boys and girls using 3, 4, and 5 degrees of freedom. The primary analysis utilized 4 degrees of freedom. Results demonstrate that the estimates remain directionally consistent despite minor numerical sensitivity to the spline specification.

**Supplementary Figure S1. Model diagnostics and sensitivity analysis.**

Top left: residuals versus fitted HAZ for the median (τ = 0.50) linear quantile mixed model, by sex. Top right: normal QQ plot of the same residuals. Bottom: comparison of the BMIZ coefficient (with 95% confidence interval (CI)) at each headline quantile, estimated three ways, the primary/secondary cluster-bootstrap linear quantile mixed model, a quantile generalized additive model (qgam), and a naive quantile regression that ignores within-child clustering, separately for boys, girls, and the pooled cohort. LQMM= Linear quantile mixed model (primary model separately for boys and girls, secondary model for pooled boys and girls).

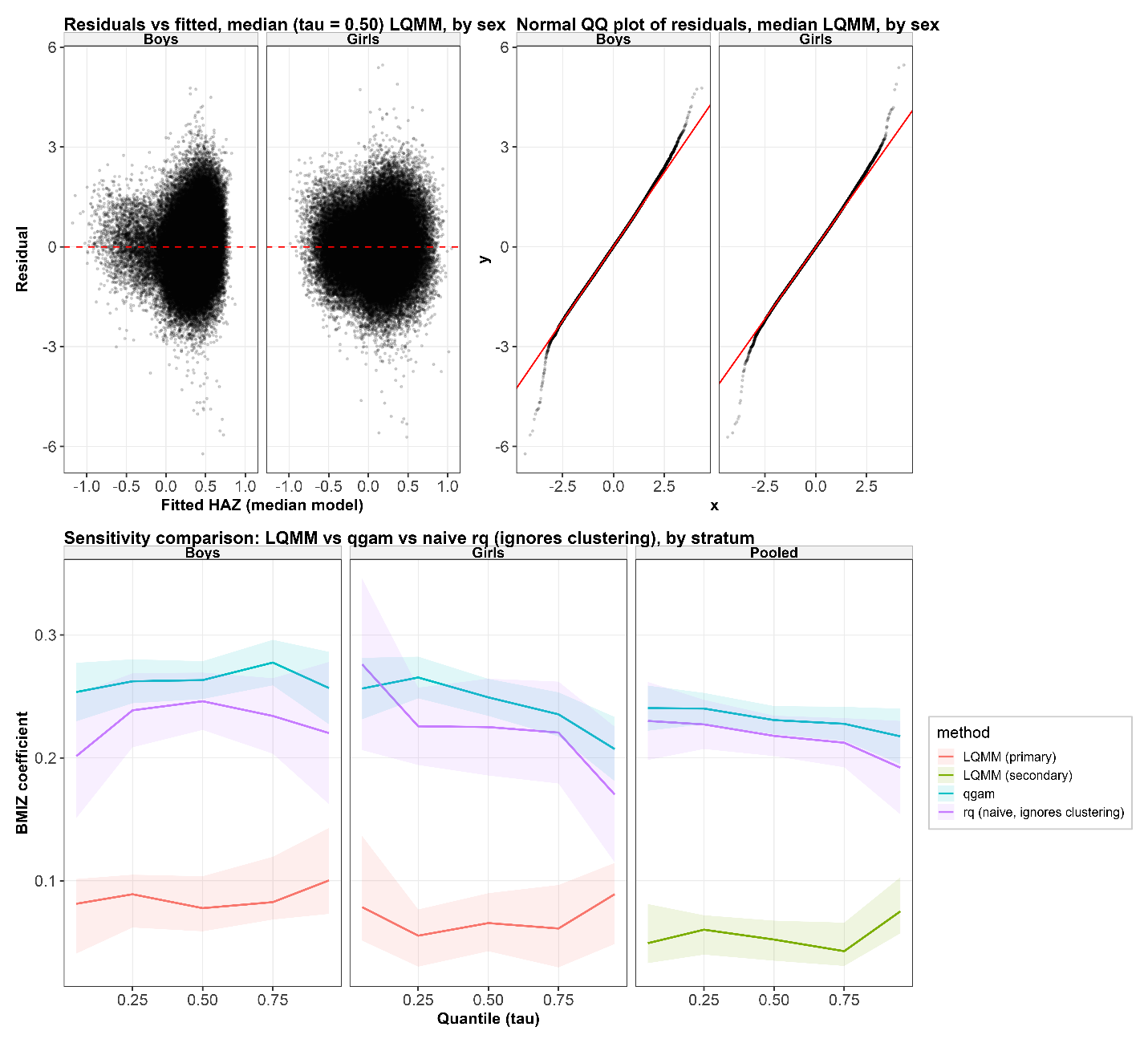
